# Routine Chest CTA Reliably Quantifies Left Ventricular Myocardial Mass

**DOI:** 10.64898/2026.10.26.26366462

**Authors:** Kelly C. Twohig, David Jiang, Joseph A. Pugar, Jeremy Slivnick, Luis Landeras, Luka Pocivavsek

## Abstract

**Background:** Routine chest computed tomography angiography (CTA) is the primary imaging modality for thoracic aortic disease, yet cardiac information within these examinations remains underutilized. We evaluated whether left ventricular myocardial mass (LVMM) can be reliably quantified from routine non–electrocardiographically (ECG)-gated chest CTA and its application in thoracic aortic pathology.

**Methods:** CTA scans were obtained from 257 patients without aortic disease and 181 patients with thoracic aortic aneurysm or dissection. LVMM was quantified using automated three-dimensional segmentation. In a separate 10-patient validation cohort, LVMM was compared across end-systolic, end-diastolic, and non-ECG-gated source reconstructions. CTA-derived LVMM was compared with published echocardiographic and cardiovascular magnetic resonance (CMR) reference values using Cohen’s d.

**Results:** LVMM remained stable across cardiac phases and non-ECG-gated images (mean coefficient of variation, 2.4%), whereas left ventricular volume varied substantially (36.9%). CTA-derived LVMM demonstrated small effect sizes with the Penn method (*d* = 0.22) and moderate effect sizes with ASE linear (*d* = 0.67), area-length (*d* = 0.68), and CMR (*d* = 0.63). LVMM increased through young adulthood, with minimal association with age beyond 30 years. LVMM was comparable between patients with thoracic aneurysms and normal adults (140 ± 43 g vs. 141 ± 40 g; adjusted *p* = 1.0) but significantly greater in acute (199 ± 61 g) and chronic (191 ± 57 g) aortic dissection (both adjusted *p* < 0.001).

**Conclusions:** Routine non-ECG-gated chest CTA enables reliable automated LVMM quantification without additional imaging or radiation and may facilitate large-scale investigations of ventricular remodeling in thoracic aortic disease.

## Introduction

For over two decades, computed tomography angiography (CTA) has served as the first-line modality for the diagnosis and longitudinal surveillance of thoracic aortic pathology. Because of its high spatial resolution, widespread availability, rapid acquisition, and reproducibility, CTA has become the cornerstone for acute evaluation, pre-operative planning, and long-term monitoring of aortic dissections, aneurysms, and other complex aortic diseases.

Despite its extensive utilization, each CTA represents a substantially underleveraged dataset. Beyond aortic diameter (long considered the gold standard for characterizing aortic disease), routine chest CTA captures comprehensive three-dimensional information regarding both vascular and cardiac morphology. Our prior work has demonstrated that detailed geometric measurements of aortic shape can be extracted from routine CTA to characterize disease beyond diameter alone [1]. In this study, we extend this framework to the heart, demonstrating that quantitative left ventricular myocardial mass (LVMM) can be derived from a routine, non-electrocardiographically (ECG)-gated chest CTA. The ability to quantify LVMM from routine CTA creates an opportunity to evaluate cardiac remodeling in patients undergoing treatment for thoracic aortic disease using imaging that is already obtained as part of standard care.

Thoracic endovascular aortic repair (TEVAR) has become increasingly prevalent in the treatment of thoracic aortic pathology. However, endovascular stent grafts are inherently stiffer and less compliant than the native aorta, raising concern for elevated left ventricular afterload and subsequent ventricular remodeling [2,3]. The cardiovascular implications of TEVAR have therefore emerged as an area of growing interest, particularly as newer-generation devices permit more proximal deployment within the aortic arch and ascending aorta. Small retrospective studies have suggested that TEVAR may be associated with increases in left ventricular mass following implantation [4,5]. However, the long-term cardiovascular consequences of TEVAR remain incompletely understood. Large-scale investigations are challenging, as vascular surgery patients are not routinely followed longitudinally with dedicated cardiac imaging modalities, such as echocardiography, ECG-gated CTA, or cardiac magnetic resonance imaging (CMR).

Leveraging routine chest CTA for quantitative assessment of left ventricular morphology before and after TEVAR offers an opportunity for comprehensive cardiovascular phenotyping without additional imaging, cost, or patient burden. In this study, we demonstrate that quantitative LVMM can be derived from routine, non–ECG-gated CTA using automated three-dimensional segmentation. We validate this approach against established echocardiographic and CMR reference standards and define age-and sex-associated variation within a large all-comer trauma population without aortic pathology. This framework establishes CTA-derived LVMM as a clinically accessible and scalable tool for large-scale investigations of ventricular remodeling. We further apply this framework to patients with thoracic aortic pathology, evaluating LVMM in aneurysms and acute/chronic dissections to characterize ventricular remodeling prior to endovascular intervention.

## Methods

### Normative cohort

The normative cohort consisted of 270 patients without aortic pathology who were originally identified for prior investigations of aortic morphology. Patients were identified through retrospective review of individuals presenting to the University of Chicago Trauma Center and Comer Children’s Hospital Trauma Center between January 1, 2010, and December 31, 2023. Trauma patients routinely undergo chest CTA, providing high-quality vascular imaging in individuals without known aortic disease.

Patients with aortic pathology, prior aortic intervention, or known connective tissue disorder were excluded. Because the cohort was defined by the absence of aortic pathology, patients were not specifically screened for preexisting myocardial disease. Patients were selected to achieve representation across sex and age groups. When multiple CTA examinations were available for the same patient during a single admission, each scan was analyzed, and measurements from repeat examinations were averaged to generate a single patient-level value for subsequent cohort analyses.

### TEVAR cohort

A separate, pre-established database of patients who underwent TEVAR for thoracic aortic pathology was used for analysis. Indications for TEVAR included acute type B aortic dissection, chronic dissection, and thoracic aortic aneurysm. CTA scans were obtained from the University of Chicago, Endeavor Health Evanston Hospital, and deidentified imaging datasets provided by medical device companies under data use agreements. Only pre-TEVAR scans were included in this study.

### Validation cohort

Because CTA scans in the primary dataset were not ECG-gated, a separate validation cohort was identified. This cohort consisted of 10 patients who underwent clinically indicated ECG-gated CTA. These patients were not included in the normative cohort. For each patient, left ventricular volume (LVV) and LVMM were measured at end-systole, end-diastole, and from the non–ECG-gated source reconstruction obtained during the same imaging session. Because the source reconstruction represents acquisition at an arbitrary point in the cardiac cycle, it approximates the non–ECG-gated scans used in the primary study cohort.

All imaging data were received in deidentified Digital Imaging and Communications in Medicine (DICOM) format. Third-party DICOM files were provided under data use agreements between the University of Chicago and Medtronic plc (Dublin, Ireland) and W. L. Gore & Associates (Newark, DE). All data collection and analyses were conducted in accordance with the Declaration of Helsinki and were approved by the University of Chicago Institutional Review Board (IRB20-0653, IRB21-0299).

### Segmentation and LVMM quantification

Three-dimensional models of the heart were generated from CTA scans using commercial software (Simpleware; Synopsys, Inc., Sunnyvale, CA, USA). Segmentations were generated automatically using built-in tools within the software. Papillary muscles were included within the myocardial segmentation when automatically classified as myocardium by the software; smaller papillary structures not captured by the algorithm were incorporated within the left ventricular cavity volume. All segmentations were visually reviewed by a single investigator for overall accuracy. Segmentations were excluded if the automated mask demonstrated substantial defects (e.g., large discontinuities, incomplete myocardial contouring, or failure of cardiac boundary definition) that precluded reliable volumetric analysis. Manual corrections were not performed.

Myocardial volume and LVV were calculated using voxel-based summation algorithms within the software. LVMM was derived by multiplying myocardial volume by a myocardial tissue density of 1.055 g/mL, a value commonly applied in human imaging studies [6].

### Validation of LVMM across the cardiac cycle

To confirm that LVMM remains stable across the cardiac cycle and that non–ECG-gated source reconstructions provide comparable measurements, validation analyses were performed in the 10-patient gated cohort. Within-patient mean, standard deviation (SD), and coefficient of variation (CV; SD divided by mean) were calculated to quantify variability in LVMM and LVV across systolic, diastolic, and source reconstructions. Pairwise comparisons of LVMM and LVV were performed using paired t-tests with Bonferroni correction for multiple comparisons. A two-sided adjusted p-value <0.01 was considered statistically significant.

To further contextualize the CTA-derived LVMM values, measurements from the normative cohort were compared with published reference values from large-scale imaging studies. Echocardiographic reference values were obtained from Lee et al. [7], which reported LVMM in 1,854 adults using five measurement techniques. Cardiovascular magnetic resonance (CMR) reference values were obtained from Maceira et al. [8], which evaluated 120 healthy adults. Because the echocardiography cohort was substantially larger than the CMR and CTA cohorts, small differences could achieve statistical significance despite a limited magnitude of effect. Therefore, Cohen’s *d* was calculated to quantify the standardized magnitude of between-group differences.

### Statistical analysis

To characterize age-related variation in LVMM within the normative cohort, the relationship between LVMM and age was evaluated using piecewise linear regression. Candidate age breakpoints were evaluated, and the breakpoint yielding the lowest sum of squared errors was selected as the optimal breakpoint. Separate linear regression models were subsequently fit above and below the identified breakpoint and stratified by sex. Model fit was assessed using the coefficient of determination (R²) and root mean square error (RMSE).

Pairwise comparisons of LVMM between diagnostic cohorts were performed using the Mann–Whitney U test with Bonferroni correction for multiple comparisons. A two-sided adjusted p-value <0.01 was considered statistically significant.

## Results

### Patient characteristics

The normative cohort initially targeted 270 patients, evenly distributed by sex (135 male and 135 female) and across predefined age groups. Following automated segmentation and quality review, 257 patients (130 male and 127 female) were included in the final analysis (**Table 1**). The majority of patients in the normative cohort were Black (70.4%). The pathologic cohort comprised 84 patients with aortic aneurysms, 34 with chronic aortic dissections, and 63 with acute aortic dissections.

**Table 1.** Demographic characteristics of the normal patient cohort, stratified by age group and sex. Black patients comprised 70.4% of the cohort; the remaining patients were White (19.1%), more than one race (7.4%), Asian/Mideast Indian (0.4%), or unknown/not reported (2.7%).

| Normal Patients |  |  |  |  |
| --- | --- | --- | --- | --- |
| Age Group (Years) | Male |  | Female |  |
|  | n | % Black | n | % Black |
| 0 - 19 | 19 | 47.4 | 18 | 72.2 |
| 20 - 29 | 15 | 86.7 | 14 | 100 |
| 30 - 39 | 14 | 71.4 | 14 | 100 |
| 40 - 49 | 15 | 80.0 | 14 | 85.7 |
| 50 - 59 | 15 | 66.7 | 14 | 85.7 |
| 60 - 69 | 13 | 61.5 | 14 | 64.3 |
| 70 - 79 | 14 | 71.4 | 14 | 64.3 |
| 80 - 89 | 15 | 46.7 | 15 | 60.0 |
| 90 - 99 | 10 | 30.0 | 10 | 70.0 |
| **Total** | **130** | **63.1%** | **127** | **78.0%** |

### Validation of myocardial mass conservation across the cardiac cycle

Three-dimensional segmentations of the left ventricular myocardium and cavity for the 10 validation patients are shown in **Figure 1**. LVMM did not differ significantly between end-systole and end-diastole (adjusted p = 0.41), with mean LVMM values of 183 g and 186 g, respectively (**Table 2)**. LVMM derived from the source (non-ECG-gated) scan was numerically lower (mean 178 g) than measurements obtained from ECG-gated phases; however, these differences did not meet the predefined threshold for statistical significance (vs. end-systole: adjusted p = 0.22; vs. end-diastole: adjusted p = 0.03). The coefficient of variation for LVMM ranged from 0.7% to 7.7%, with a mean of 2.4%.

**Figure 1.**
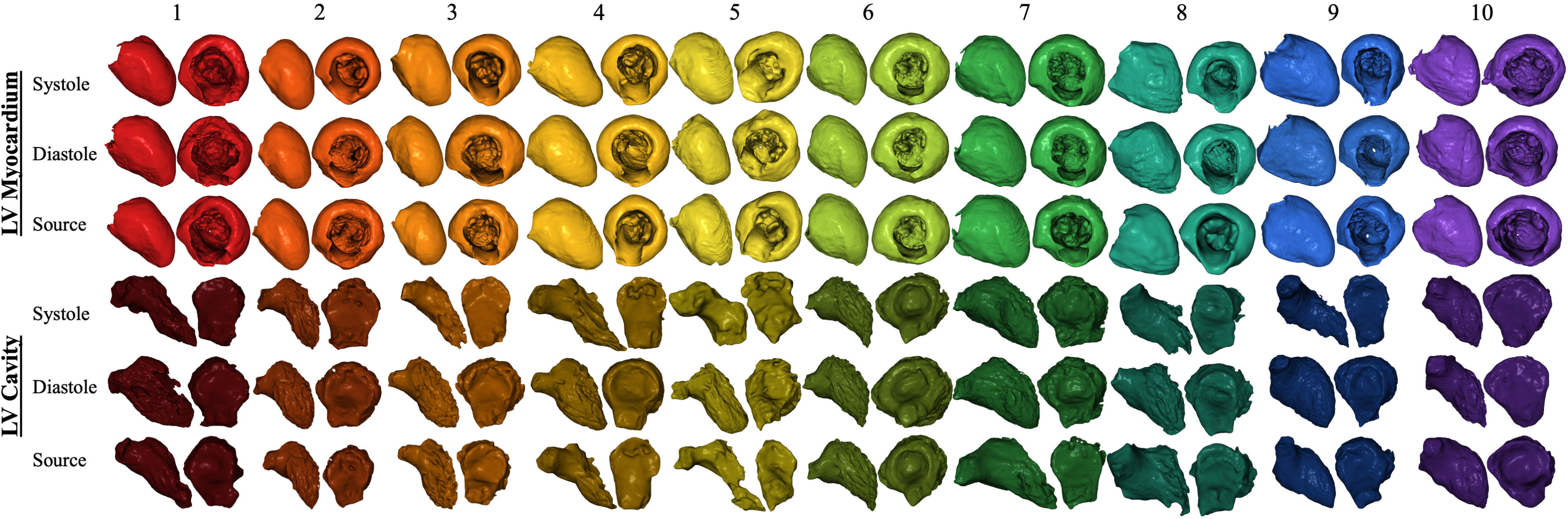
Three-dimensional segmentations used to validate conservation of left ventricular myocardial mass (LVMM) throughout the cardiac cycle. Ten patients underwent ECG-gated CT and CTA source scans in immediate succession. The left ventricular myocardium (top) and cavity (bottom) are shown for each patient at end-systole, end-diastole, and on the CTA source scan. Each patient is represented by a unique color, with darker shades denoting the corresponding left ventricular cavity.

**Table 2.** Quantitative validation of left ventricular myocardial mass (LVMM) conservation throughout the cardiac cycle. LVMM and left ventricular volume (LVV) were measured at end-systole, end-diastole, and from the CTA source scan for each of the 10 validation patients. LVMM did not differ significantly across measurements (adjusted paired *t*-test: end-systole vs. end-diastole, *p* = 0.41; end-systole vs. source, *p* = 0.22; end-diastole vs. source, *p* = 0.03), whereas LVV differed significantly between end-systole and end-diastole (*p* < 0.001) and between end-diastole and the source scan (*p* = 0.003). Coefficients of variation were calculated from unrounded values; displayed values are rounded for presentation.

| Left Ventricular Myocardial Mass (LVMM) (g) |  |  |  |  |  |
| --- | --- | --- | --- | --- | --- |
| Patient Number | End-Systole | End-Diastole | Source | Average (± SD) | Coefficient of Variation (%) |
| 1 | 133 | 134 | 135 | 134 ± 1 | 0.7 |
| 2 | 220 | 226 | 213 | 219 ± 6 | 2.9 |
| 3 | 234 | 248 | 226 | 236 ± 11 | 4.7 |
| 4 | 148 | 152 | 141 | 147 ± 6 | 3.8 |
| 5 | 119 | 125 | 122 | 122 ± 3 | 2.8 |
| 6 | 242 | 239 | 232 | 238 ± 5 | 2.2 |
| 7 | 304 | 304 | 296 | 301 ± 5 | 1.5 |
| 8 | 127 | 126 | 130 | 128 ± 2 | 1.9 |
| 9 | 181 | 175 | 155 | 170 ± 13 | 7.7 |
| 10 | 124 | 133 | 124 | 127 ± 5 | 4.1 |
| Average | 183 | 186 | 178 | 182 ± 4 | 2.4 |

**Table 2.** Quantitative validation of left ventricular myocardial mass (LVMM) conservation throughout the cardiac cycle.
| Left Ventricular Volume (cc) |  |  |  |  |  |
| --- | --- | --- | --- | --- | --- |
| Patient Number | End-Systole | End-Diastole | Source | Average (± SD) | Coefficient of Variation (%) |
| 1 | 42 | 101 | 74 | 72 ± 30 | 41.0 |
| 2 | 96 | 182 | 137 | 138 ± 43 | 31.1 |
| 3 | 61 | 173 | 119 | 118 ± 56 | 47.4 |
| 4 | 43 | 125 | 43 | 71 ± 47 | 67.1 |
| 5 | 12 | 82 | 20 | 38 ± 38 | 99.5 |
| 6 | 124 | 165 | 133 | 141 ± 21 | 15.2 |
| 7 | 169 | 202 | 88 | 153 ± 59 | 38.2 |
| 8 | 26 | 81 | 43 | 50 ± 28 | 56.3 |
| 9 | 56 | 176 | 135 | 122 ± 61 | 49.9 |
| 10 | 37 | 76 | 81 | 65 ± 24 | 37.3 |
| Average | 67 | 136 | 87 | 97 ± 36 | 36.9 |

In contrast, LVV varied significantly across the cardiac cycle (adjusted p < 0.001) with mean volumes of 67 mL at end-systole and 136 mL at end-diastole. LVV derived from the source scan (mean 87 mL) was intermediate between systolic and diastolic measurements. The coefficient of variation for LVV ranged from 15.2% to 99.5%, with a mean of 36.9%.

### Comparison of LVMM calculations by various methods

**Table 3** compares LVMM derived from normative CTA with values reported using echocardiography (Lee et al. [7]) and CMR (Maceira et al. [8]). Among male patients, mean CTA-derived LVMM (156 ± 40 g) was similar to values reported using the Penn-cube technique (152 ± 46 g) and CMR (146 ± 20 g). Among female patients, echocardiographic measurements generally demonstrated a progressive age-related increase in LVMM, whereas CTA-derived measurements did not demonstrate a consistent age-related trend. Across the overall adult cohort, mean CTA-derived LVMM (141 ± 40 g) was higher than values reported using each of the five echocardiographic methods and CMR.

**Table 3.**
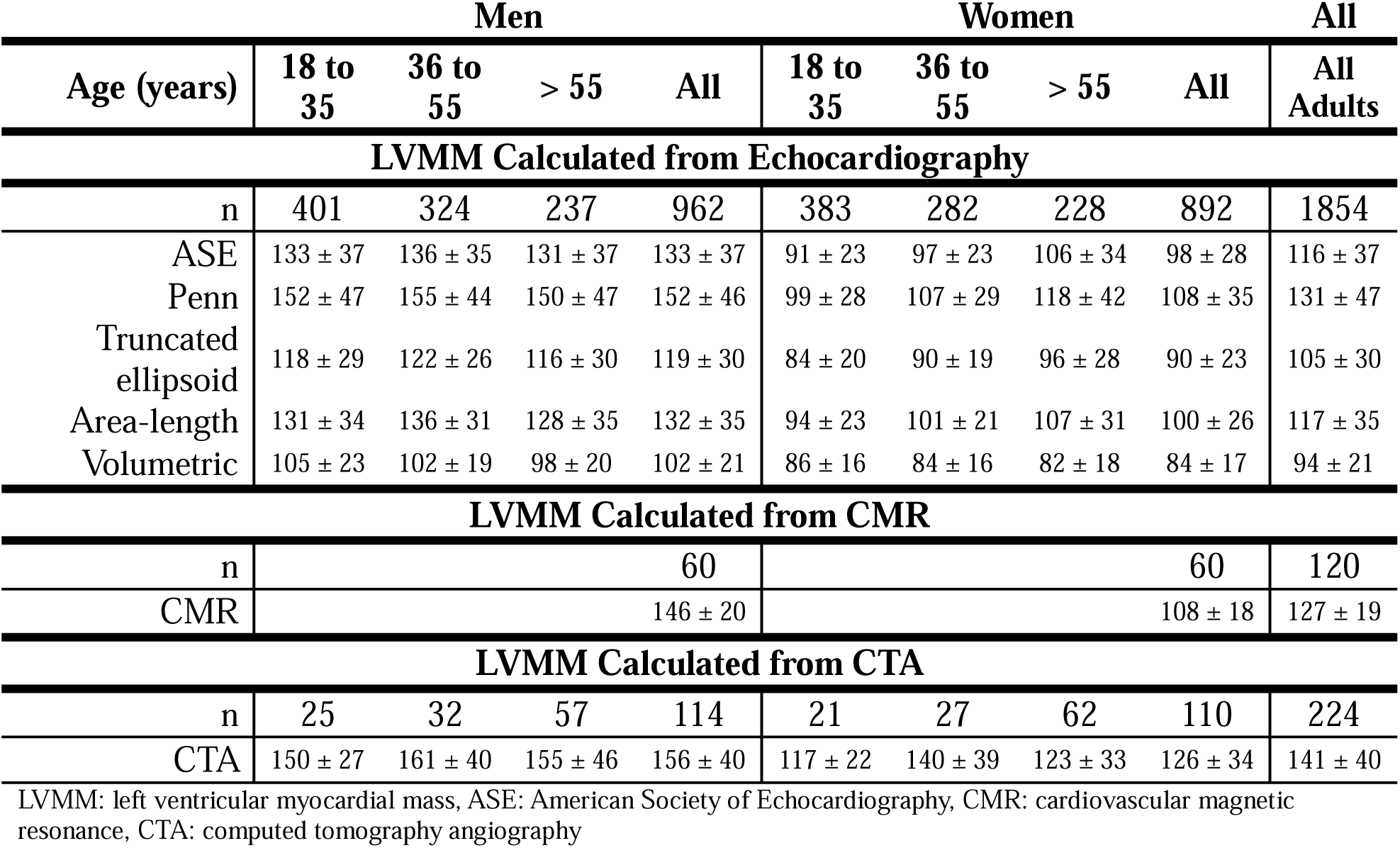
Comparison of left ventricular myocardial mass (LVMM) derived from non-ECG-gated CTA with published reference values derived from echocardiography (Lee *et al* [7]) and cardiovascular magnetic resonance (CMR; Maceira et al. [8]). Values are reported as mean LVMM (g) ± standard deviation.

**Figure 2** presents Cohen’s *d* effect sizes comparing CTA-derived LVMM with values obtained using echocardiographic methods and CMR. By convention, *d* values of 0.2, 0.5, and 0.8 represent small, medium, and large effect sizes, respectively. Among adults, the effect size was small for the Penn-cube method (*d* = 0.22); medium for the ASE linear (*d* = 0.67) and area-length (*d* = 0.68) methods and CMR (*d* = 0.63); and large for the truncated ellipsoid (*d* = 1.15) and volumetric (*d* = 1.98) methods.

**Figure 2.**
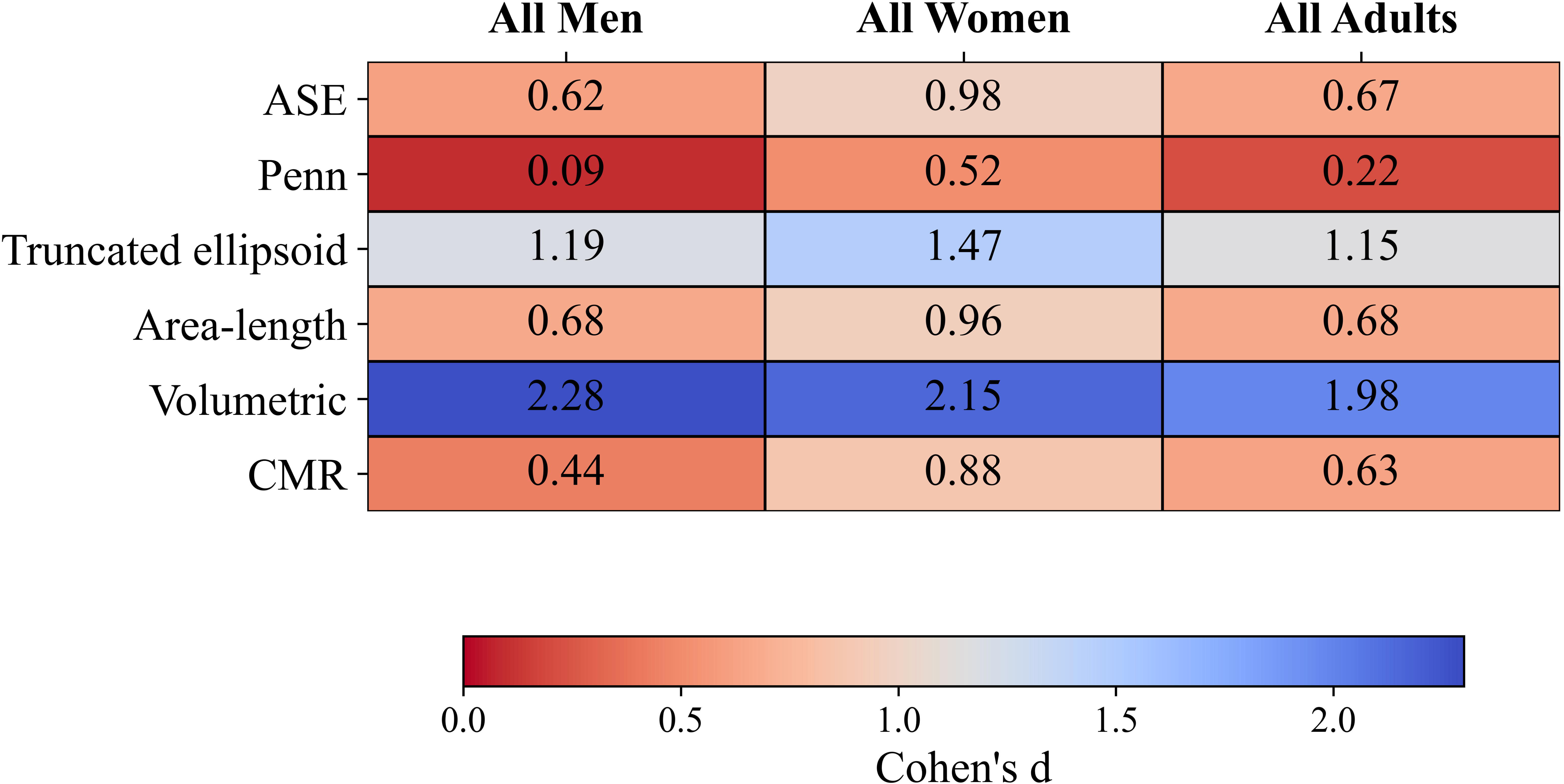
Comparison of CTA-derived left ventricular myocardial mass (LVMM) with published reference values obtained using linear (ASE and Penn), two-dimensional (truncated ellipsoid and area-length), and three-dimensional (volumetric) echocardiographic techniques, as well as cardiovascular magnetic resonance (CMR). Values represent the absolute Cohen’s d effect size, calculated using the sample-size-weighted pooled standard deviation. Lower values (red) indicate greater similarity to CTA-derived LVMM, whereas higher values (blue) indicate greater differences.

### Age-related variation in LVMM

**Panels A-C** (**Figure 3)** illustrate LVMM as a function of age in the normative cohort. Piecewise regression identified 30 years as the optimal age breakpoint, corresponding to the lowest sum of squared errors. LVMM increased rapidly between 2 and 30 years of age, at a rate of 3.98 g/year (**Panel C**). Beyond 30 years, LVMM demonstrated minimal association with age (**Panel C**, *R*^2^ = 0.02).

**Figure 3.**
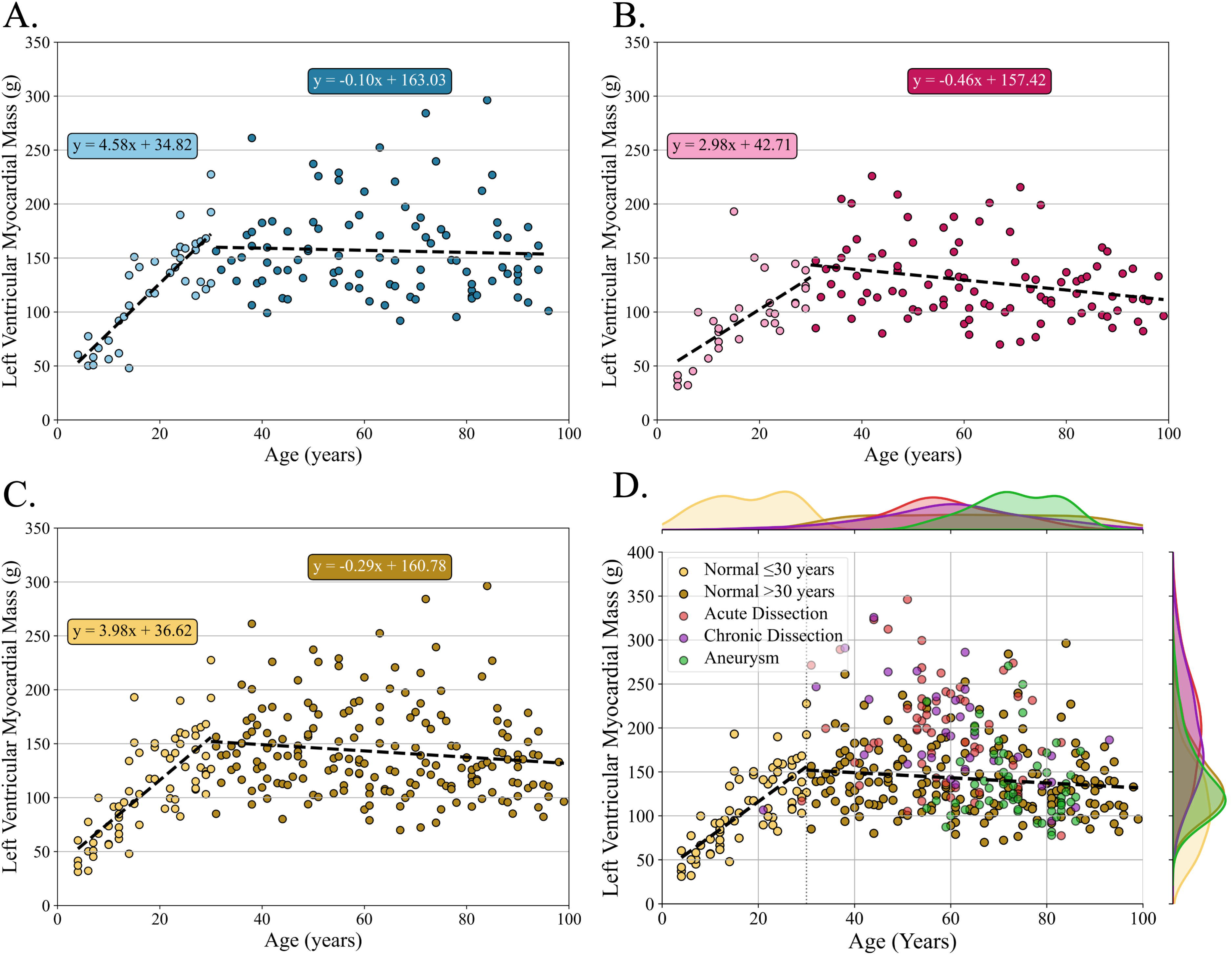
Left ventricular myocardial mass (LVMM) plotted by age. Normal patients are shown stratified by male sex (A), female sex (B), and overall (C); patients with acute dissection, chronic dissection, and aneurysm are shown in (D). Age 30 years was selected as the breakpoint based on the lowest sum of squared errors. Best-fit line metrics for the normal cohort were as follows: (A) ≤30 years, R² = 0.68, RMSE = 25.48 g; >30 years, R² = 0.002, RMSE = 42.34 g; (B) ≤30 years, R² = 0.44, RMSE = 26.98 g; >30 years, R² = 0.067, RMSE = 33.61 g; and (C) ≤30 years, R² = 0.56, RMSE = 28.90 g; >30 years, R² = 0.02, RMSE = 40.97 g.

**Panel D** incorporates patients with aortic aneurysms and dissections prior to undergoing TEVAR. The right axis displays the one-dimensional LVMM distributions for each diagnostic cohort. LVMM values among patients with aortic aneurysms demonstrated substantial overlap with the normative cohort. In contrast, the acute and chronic dissection cohorts demonstrated broader LVMM distributions, with some patients overlapping the normative range and others exhibiting substantially elevated LVMM.

**Figure 4** presents LVMM distributions across diagnostic cohorts using violin plots. Mean LVMM among patients with thoracic aortic aneurysms (140 ± 43 g) did not differ significantly from that of normal adults (141 ± 40 g; adjusted *p* = 1). In contrast, patients with acute dissections (199 ± 61 g) and chronic dissections (191 ± 57 g) demonstrated significantly higher LVMM than normal adults (adjusted *p* < 0.001 for both comparisons). The violin plots further illustrate the heterogeneity of LVMM within the dissection cohorts noted in **Figure 3**.

**Figure 4.**
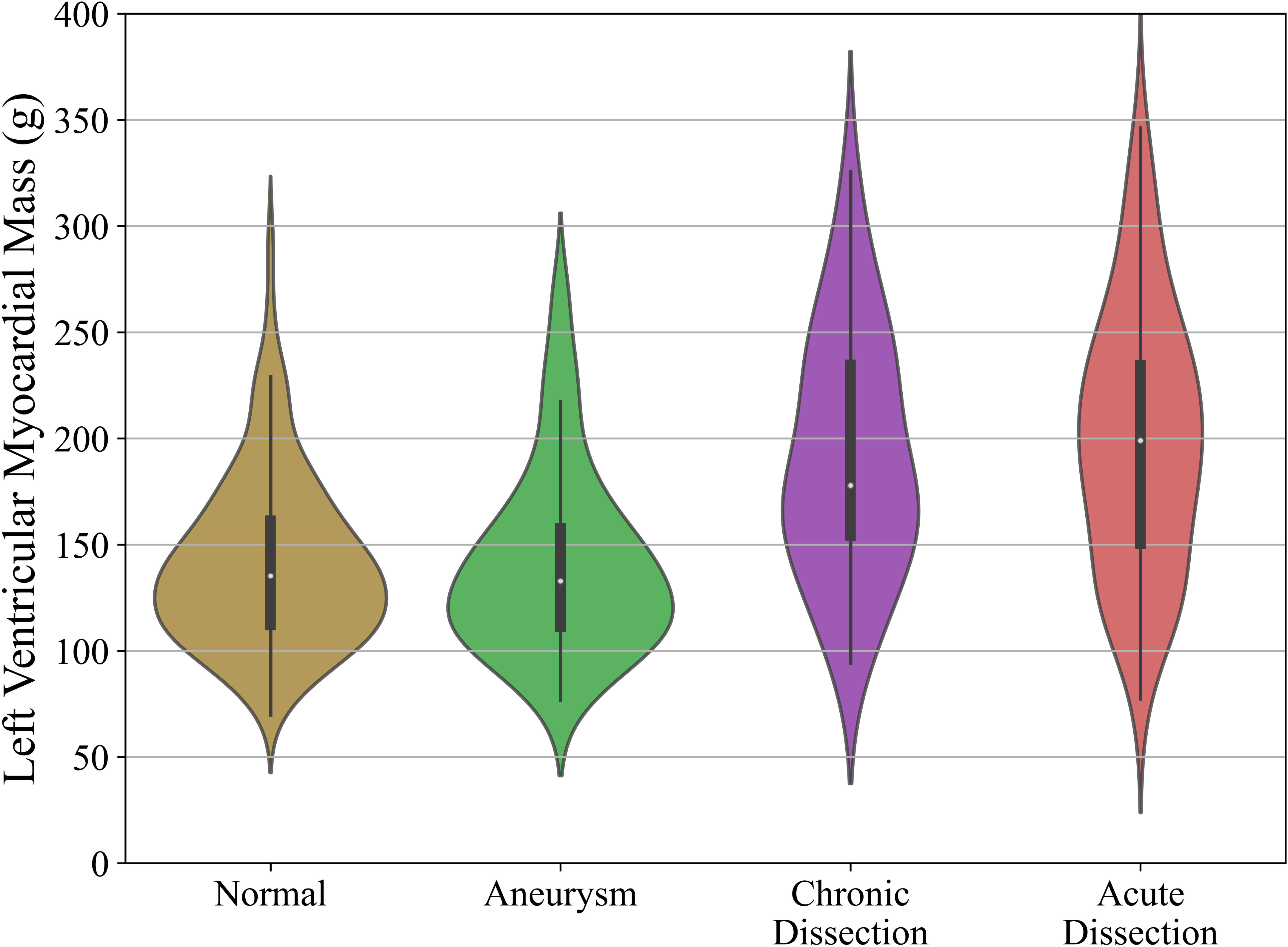
Violin plots illustrating the distribution of left ventricular myocardial mass (LVMM) among normal adults and patients with thoracic aortic aneurysm, chronic aortic dissection, or acute aortic dissection. LVMM was higher in the acute and chronic dissection cohorts compared with normal adults, whereas the aneurysm cohort demonstrated a similar distribution to the normal cohort. The dissection cohorts demonstrated broader distributions with visually apparent bimodality. Central markers represent mean LVMM, and the width of each violin reflects data density across the observed range.

## Discussion

In this study, we demonstrate that LVMM can be reliably quantified from routine non-ECG-gated CTA. LVMM remained stable across the cardiac cycle and demonstrated reasonable agreement with published reference values derived from echocardiography and CMR[7,8]. In addition to validating this measurement approach, we observed significantly higher LVMM among patients with acute and chronic aortic dissections compared with normal adults. Finally, the substantial variability in LVMM within the dissection cohorts suggests underlying phenotypic heterogeneity.

Because myocardial mass is a structural property, it should remain constant across systole and diastole. Our findings support this principle, demonstrating minimal variation in LVMM across end-systolic, end-diastolic, and source reconstructions. In contrast, LVV varied substantially throughout the cardiac cycle, consistent with the expected physiologic variation in ventricular volume and its dependence on cardiac phase. Small differences between source and ECG-gated reconstructions may reflect reduced myocardial edge definition on non-gated images, potentially resulting in mild under-segmentation. Taken together, these findings support the reliability of myocardial mass quantification from routine non–ECG-gated CTA without the need for dedicated cardiac acquisition.

**Table 3** and **Figure 2** demonstrate that CTA-derived LVMM is broadly consistent with values reported in large reference cohorts using established imaging modalities, with the smallest difference observed relative to the Penn-cube echocardiographic method. CTA-derived LVMM demonstrated medium effect sizes relative to values derived using the ASE linear method, which is more commonly used in routine clinical practice, as well as the area-length method and CMR. In contrast, larger differences were observed relative to the truncated ellipsoid and volumetric echocardiographic methods.

Average LVMM measured by CTA was modestly higher than values reported across echocardiographic techniques and CMR, with this difference more pronounced among women. One potential explanation is the inclusion of papillary muscle within the automated myocardial segmentation, which may proportionally increase measured mass to a greater extent in smaller ventricles. Differences in cohort composition may also contribute to this discordance. Our normative cohort represents an all-comer clinical population with a high proportion of Black patients and potentially broader cardiovascular risk profiles than the carefully selected healthy reference populations used in many echocardiographic and CMR studies. Population-based investigations have demonstrated higher average left ventricular mass and a greater prevalence of left ventricular hypertrophy among Black individuals, even after adjustment for body composition and blood pressure [9–11]. These demographic and methodological differences may partially account for the higher LVMM observed in our cohort.

These observations highlight the importance of establishing modality-specific reference standards. LVMM obtained from CTA should ideally be interpreted relative to reference values derived using similar acquisition and segmentation methodologies. Further investigation is needed to establish robust normative ranges for CTA-derived LVMM, with adequate representation across age, sex, and racial groups to reflect the diversity of contemporary patient populations. Rather than replacing established imaging modalities, CTA-derived LVMM may serve as a complementary metric, expanding the cardiovascular information obtainable from routine thoracic imaging.

Beyond comparisons with published reference values, CTA-derived LVMM also reproduced established age-related patterns in myocardial mass. In this cohort, which included pediatric patients, LVMM increased rapidly during childhood and young adulthood, consistent with somatic growth and physiologic myocardial remodeling during maturation [12,13]. Beyond 30 years of age, LVMM demonstrated minimal association with age, consistent with prior CMR-based investigations demonstrating relative stability of myocardial mass after maturation [14–16]. The ability of CTA-derived LVMM to reproduce these well-described biologic trends provides additional support for the construct validity of this measurement approach.

The ability to readily quantify LVMM in patients with thoracic aortic pathology carries important clinical implications. In cardiovascular literature, myocardial mass is widely used as a marker of ventricular remodeling and cumulative hemodynamic burden [17]. Increased LVMM is associated with chronic hypertension, concentric remodeling, and adverse cardiovascular outcomes, including heart failure and mortality [18–20]. CTA-derived LVMM therefore offers an opportunity to characterize ventricular remodeling at the time of aortic diagnosis. In patients undergoing endovascular repair, LVMM may serve as a noninvasive marker of systemic cardiovascular phenotype, enabling assessment of preoperative remodeling and longitudinal evaluation following intervention.

We further demonstrate significantly elevated LVMM in patients presenting with acute and chronic aortic dissections. While consistent with the known association between aortic dissection and chronic hypertension [21,22], the presence of ventricular heterogeneity within this cohort is particularly notable. Some patients exhibited markedly elevated LVMM, suggesting variable degrees of myocardial remodeling among patients with dissection.

Because LVMM reflects cumulative pressure overload and hypertensive remodeling, elevated values may represent greater systemic end-organ involvement and could have implications for perioperative cardiovascular risk. Although exploratory, these findings raise the possibility that LVMM may aid in risk stratification among patients undergoing endovascular repair. Identification of individuals with markedly elevated LVMM may support more aggressive blood pressure optimization, heightened perioperative hemodynamic monitoring, and closer longitudinal surveillance following intervention. Prospective studies evaluating longitudinal changes in LVMM after repair and their association with clinical outcomes are warranted.

This study has several limitations. It is retrospective in design. Segmentation was automated and visually inspected but not manually refined, and inclusion of papillary muscle within the myocardial compartment may have influenced measured mass. Direct head-to-head comparison with echocardiography or CMR in the same patients was not performed.

Additionally, LVMM was evaluated as absolute mass rather than indexed to body surface area because reliable measured height and weight were not consistently available, particularly within the normal cohort. Accordingly, differences in body size between cohorts may contribute to observed differences in LVMM. Finally, longitudinal clinical outcomes were not consistently available, precluding assessment of whether elevated CTA-derived LVMM is associated with adverse cardiovascular or aortic outcomes. Therefore, this study establishes the feasibility and reliability of CTA-derived LVMM quantification but does not establish its prognostic significance. Future studies linking longitudinal changes in LVMM with clinical outcomes will be necessary to determine the clinical utility of this imaging biomarker.

## Conclusion

LVMM can be reliably derived from routine non–ECG-gated CTA and demonstrates physiologic validity across the cardiac cycle and lifespan. Elevated and heterogeneous LVMM in patients with aortic dissection suggests that routine CTA may provide insight into systemic cardiovascular remodeling beyond aortic morphology alone.

## Data Availability

All data produced in the present study are available upon reasonable request to the authors

## Abbreviations

ASE: American Society of Echocardiography
CTA: computed tomography angiography
CMR: cardiovascular magnetic resonance
LVMM: left ventricular myocardial mass
LVV: left ventricular volume
TEVAR: thoracic endovascular aortic repair

